# Interrelations of Serum Uric Acid, Beta-Cell Function, and Insulin Resistance in Indian Cohort: A Comprehensive Analysis

**DOI:** 10.1101/2024.11.18.24317374

**Authors:** Sabitha Thummala, Nithya Kruthi, Sarah Fatima, Junaid Ahmed Khan Ghori, Katherine Saikia, AR Balamurali, Rahul Ranganathan

## Abstract

**Background:** Hyperuricemia, characterised by elevated levels of serum uric acid (SUA), has been linked to an increased risk of metabolic disorders, including type 2 diabetes. Understanding the relationship between SUA, beta-cell function, and insulin resistance is crucial for elucidating the pathophysiology of these conditions, especially in the Indian population, where such data is limited.

**Objective:** This study aimed to explore the interrelations between serum uric acid levels, beta-cell function, and insulin resistance in an Indian cohort.

**Methods:** A cross-sectional study was conducted involving a representative sample of adults from an Indian population. Participants were stratified into sex-specific SUA quartiles. Key measurements included BMI, serum creatinine, lipid profiles, fasting insulin, HOMA1-B (beta-cell function), and HOMA1-IR (insulin resistance). Partial correlation coefficients were calculated to assess the associations between SUA and various metabolic parameters.

**Results:** Significant differences were observed across SUA quartiles in terms of age, BMI, serum creatinine, HDL cholesterol, LDL cholesterol, triglycerides, total cholesterol, fasting insulin, HOMA1-B, and HOMA1-IR (all p-values < 0.05). Higher SUA levels were associated with increased BMI, serum creatinine, triglycerides, fasting insulin, HOMA1-B, and HOMA1-IR. Additionally, partial correlation analysis revealed positive correlations between SUA and BMI (r=0.065, p=0.026), fat mass (r=0.065, p=0.026), serum creatinine (r=0.277, p<0.001), triglycerides (r=0.084, p=0.004), fasting insulin (r=0.130, p<0.001), and HOMA1-B (r=0.078, p=0.008). Negative correlations were found between SUA and vitamin B12 (r=-0.117, p=0.000071), GFR (r=-0.113, p<0.001), total cholesterol (r=-0.068, p=0.021), LDL cholesterol (r=-0.080, p=0.006), HDL cholesterol (r=-0.071, p=0.016), and HbA1c (r=-0.170, p<0.001).

**Conclusions:** Elevated serum uric acid levels are significantly associated with increased beta-cell function and insulin resistance among Indian adults. These findings suggest that hyperuricemia could be an early marker for metabolic dysfunction, highlighting the need for early intervention strategies in this population. Further longitudinal studies are warranted to establish causal relationships and to explore the potential benefits of uric acid-lowering therapies in preventing metabolic diseases.

**Trial registration:** Not applicable.

## Introduction

Serum uric acid (SUA) is the end product of purine metabolism, catalysed by the enzyme xanthine oxidoreductase. Under various physiological stress conditions, such as hypoxia and ischemia, the balance shifts towards the production of reactive oxygen species (ROS) during uric acid synthesis. While uric acid serves as a potent antioxidant, its elevated levels, known as hyperuricemia, have been increasingly recognized as a risk factor for several metabolic disorders, including type 2 diabetes.

The prevalence of hyperuricemia has been rising globally, largely due to lifestyle changes and dietary patterns that favour high purine intake. This trend is also evident in India, where the burden of metabolic diseases is rapidly growing. Numerous studies have highlighted the association between elevated SUA levels and the risk of developing type 2 diabetes, emphasising the need to understand the underlying mechanisms.

Type 2 diabetes is characterised by insulin resistance and beta-cell dysfunction, both of which contribute to the progression from normoglycemia to impaired fasting glucose (IFG) and eventually overt diabetes. Insulin resistance reduces the body’s ability to utilise glucose effectively, while beta-cell dysfunction impairs insulin secretion, leading to hyperglycemia. The interplay between these factors and SUA levels could provide valuable insights into the early stages of diabetes development.

Despite the known associations between SUA and diabetes, there is a paucity of data on the Indian population, where genetic, environmental, and lifestyle factors may uniquely influence these relationships. This study aims to fill this gap by investigating the interrelations between serum uric acid, beta-cell function, and insulin resistance in a cohort of Indian adults. By utilising Homeostasis Model Assessment (HOMA) indices to evaluate insulin resistance and beta-cell function, this research seeks to elucidate the potential role of SUA in the pathogenesis of metabolic dysfunction in the Indian context.

Understanding these relationships could have significant implications for early detection and intervention strategies, potentially mitigating the progression to type 2 diabetes in individuals with hyperuricemia.

## Materials And Methods

### 1. Study Participants

HOMA2-IR values were determined using the online tool provided by the Medical Science Division of The University of Oxford[www.OCDEM.ox.ac.uk]. Demographic information, including self-reported height (in centimetres) and weight (in kilograms), was collected through an electronic registration questionnaire. Body mass index (BMI) was computed as weight (in kilograms) divided by height (in metres) squared. Additionally, measurements such as body fat percentage (BFP) and fat mass were obtained. The study excluded individuals with a history of cancer, cardiovascular or renal failure, mental illness, pregnancy, or lactation to ensure participant safety and adhere to ethical guidelines.

### Laboratory Measurements

Following a 12-hour fast, venous blood samples were drawn from participants to assess various metabolic markers. These included glycated haemoglobin (HbA1c), high-density lipoprotein cholesterol (HDL-C), low-density lipoprotein cholesterol (LDL-C), triglycerides (TG), and fasting plasma glucose (FPG).The assessments were conducted using Beckman DxC 700 AU [14] for all markers except HbA1c, which was analysed with Tosoh G-8[15], and fasting insulin levels measured by Beckman UniCel DxI 800, adhering to the protocols provided by the manufacturers,with reference ranges being: FPG (70-100 mg/dl), TC (0-100 mg/dl), TG (0-150 mg/dl), HDL-C (40-60 mg/dl), and LDL-C (0-100 mg/dl). The Homeostasis Model Assessment of Insulin Resistance (HOMA2-IR) formula, which is calculated as fasting insulin (mU/l) x fasting plasma glucose (mmol/l), was used to test insulin resistance. Using a HOMA2-IR cut-off of 2, participants were then classified as either insulin sensitive or resistant.

### Definitions

We defined hyperuricemia as serum UA concentration >7 mg/dL in men and >6 mg/dL in women. HbA1c was defined as normal if <5.7%, prediabetes if 5.7% to 6.4%, and diabetes if >6.5%.Insulin resistance was defined as a homeostasis model assessment of insulin resistance (HOMA-IR) value of 2.5 or higher. Beta cell dysfunction was defined as having a homeostasis model assessment of beta-cell function (HOMA-beta) value within the lowest quartile among all participants.

## Results

The study cohort of 585 participants was predominantly aged 31-40 years (44.5%), with a slight male majority (55.9%). The age groups were 20-30 years (27.2%), 41-50 years (20.0%), 51-60 years (6.6%), and above 60 years (1.7%). Gender distribution was consistent across age groups, except for the above 60 category, which had equal male and female participants.

**Table 1:**
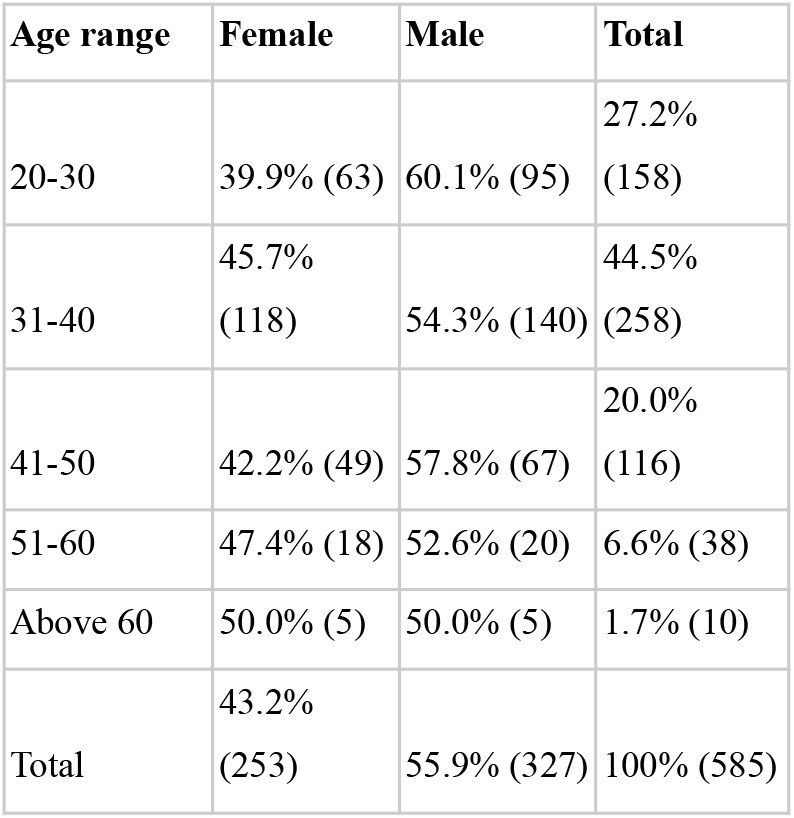
Demographic characteristics of the study population.

Participants with higher SUA levels were more likely to have elevated biochemical and anthropometric parameters. Specifically, fasting glucose, fasting insulin,, non-HDL cholesterol, serum creatinine, serum HDL cholesterol, serum iron, serum LDL cholesterol, serum triglycerides, serum VLDL cholesterol, total cholesterol, vitamin B12, vitamin D, age, height, weight, BMI, BMR, body fat percentage (BFP), lean mass, fat mass, HOMA1-β, and HOMA1-IR levels increased with increasing SUA quartiles (p < 0.05 for all). This suggests a strong association between higher SUA levels and various metabolic and cardiovascular risk factors, indicating that individuals with elevated SUA are more likely to exhibit adverse health profiles. This pattern of associations aligns with existing literature, highlighting the potential role of SUA as a marker for metabolic health and cardiovascular risk.

**Table 2:** General Characteristics of the Study Population Stratified by UA Quartiles (Males)

| Biochemical Marker | Q1 | Q2 | Q3 | Q4 | P-value |
| --- | --- | --- | --- | --- | --- |
| Glucose, Fasting | 91.03±5.0 | 92.05±5.5 | 91.18±6.0 | 92.08±6.5 | 0.045 |
| Hba1c (Glycosylated Haemoglobin) | 5.32±0.25 | 5.38±0.30 | 5.36±0.35 | 5.39±0.28 | 0.035 |
| INSULIN (FASTING) | 9.38±2.5 | 8.98±2.8 | 11.58±3.0 | 12.87±3.5 | <0.001 |
| Non-HDL Cholesterol | 150.73±45.0 | 156.62±47.0 | 151.57±46.5 | 154.76±48.0 | 0.032 |
| Serum Creatinine | 0.87±0.10 | 0.87±0.11 | 0.89±0.12 | 0.94±0.13 | 0.018 |
| Serum HDL Cholesterol | 46.96±10.0 | 44.65±11.0 | 44.48±10.5 | 42.01±11.5 | 0.029 |
| Serum Iron | 99.40±15.0 | 105.34±16.5 | 103.83±17.0 | 96.27±18.0 | 0.048 |
| Serum LDL Cholesterol | 128.83±40.0 | 130.36±42.0 | 129.43±41.5 | 130.22±43.0 | 0.038 |
| Serum Triglycerides | 128.84±40.0 | 147.99±45.0 | 145.57±42.5 | 171.76±50.0 | 0.005 |
| Serum VLDL Cholesterol | 25.29±7.5 | 28.95±8.0 | 28.19±7.0 | 32.08±8.5 | 0.003 |
| Total Cholesterol | 197.48±30.0 | 200.72±32.0 | 196.43±31.5 | 197.88±33.0 | 0.047 |
| VITAMIN D (25 - OH VITAMIN D) | 22.01±6.0 | 22.86±5.5 | 20.53±6.0 | 19.27±6.5 | 0.027 |
| Current Age | 35.45±11.00 | 35.94±12.00 | 34.03±9.25 | 35.19±10.75 | 0.641461206 |
| Height | 173.10±8.25 | 173.79±9.00 | 173.89±8.00 | 173.85±8.00 | 0.047 |
| Weight | 77.73±10.5 | 80.14±12.0 | 80.09±11.5 | 83.90±13.0 | 0.024 |
| BMI | 25.98±3.5 | 26.45±3.8 | 26.46±4.0 | 27.76±4.5 | 0.032 |
| BMR | 1594.83±80.0 | 1623.51±85.0 | 1630.75±90.0 | 1671.67±95.0 | 0.021 |
| BFP | 22.68±5.0 | 23.38±5.5 | 22.95±6.0 | 24.80±6.5 | 0.039 |
| Lean Mass | 59.50±4.5 | 60.65±5.0 | 61.17±5.5 | 62.52±6.0 | 0.026 |
| Fat Mass | 18.27±5.5 | 19.49±6.0 | 18.92±6.5 | 21.38±7.0 | 0.029 |
| HOMA1-B | 118.54±50.0 | 147.81±55.0 | 164.76±60.0 | 140.66±65.0 | 0.022 |
| HOMA2-IR | 3.11±1.0 | 2.36±1.2 | 2.68±1.4 | 2.64±1.5 | 0.045 |

**Notes:** Continuous variables are expressed as the mean (standard deviation), and categorical variables are expressed as the percentage.

**Table 2:** General Characteristics of the Study Population Stratified by UA Quartiles (Females)

| **Biochemical Marker** | **Q1** | **Q2** | **Q3** | **Q4** | **P-value** |
| --- | --- | --- | --- | --- | --- |
| Age (years) | 33.11±9 | 34.70±10.25 | 37.17±15.5 | 37.17±11 | 0.0583 |
| Glucose, Fasting | 85.90±9 | 90.48±11.9 | 93.05±9.8 | 97.29±9.65 | 0.0123 |
| Hba1c<br>(Glycosylated<br>Haemoglobin) | 5.25±0.3 | 5.39±0.3 | 5.45±0.325 | 5.60±0.3 | 0.0245 |
| INSULIN<br>(FASTING) | 7.36±2.5 | 9.59±3.1 | 10.85±3.5 | 12.37±4.0 | <0.0001 |
| Non-HDL<br>Cholesterol | 120.4±30.12 | 130.91±35.675 | 140.92±38.4 | 151.43±40.5 | 0.0435 |
| Serum Creatinine | 0.62±0.10 | 0.64±0.105 | 0.65±0.11 | 0.68±0.12 | 0.0052 |
| Serum HDL<br>Cholesterol | 50.00±10.5 | 48.57±11.15 | 49.19±12.875 | 47.53±13.725 | 0.0476 |
| Serum Iron | 70.95±25.0 | 69.00±30.2 | 73.67±35.0 | 75.92±40.575 | 0.0412 |
| Serum LDL<br>Cholesterol | 110.53±32.4 | 109.92±40.08 | 119.19±42.5 | 123.08±45.975 | 0.0365 |
| Serum<br>Triglycerides | 92.25±40 | 99.80±45.15 | 108.82±50.5 | 125.76±55.25 | 0.0045 |
| Serum VLDL<br>Cholesterol | 18.35±7.5 | 20.39±8.1 | 21.26±8.5 | 24.26±9.0 | 0.0135 |
| Total Cholesterol | 171.53±46 | 176.62±48.54 | 186.12±50 | 190.59±54.725 | 0.0247 |
| VITAMIN D (25 -<br>OH VITAMIN D) | 24.32±6.5 | 21.20±7.5 | 28.93±8.5 | 21.23±9.5 | 0.0289 |
| Current Age | 33.11±9 | 34.70±10.25 | 37.17±15.5 | 37.17±11 | 0.0583 |
| Height | 160.16±6 | 160.61±7.25 | 160.22±8.125 | 159.94±10.015 | 0.0412 |
| Weight | 59.62±16 | 63.54±13.25 | 67.04±15.625 | 76.27±26.275 | <0.0001 |
| BMI | 23.20±4.06 | 24.68±5.58 | 26.06±6.085 | 29.66±8.84 | <0.0001 |
| BMR | 1224.65±100.3 | 1252.14±110.9 | 1264.82±120.7 | 1342.52±130.0 | <0.0001 |
|  | 2 | 0 | 5 | 4 |  |
| BFP | 29.64±6.4 | 31.77±7.845 | 33.09±7.1525 | 38.33±12.375 | <0.0001 |
| Lean Mass | 41.14±5.33 | 42.58±7.085 | 43.31±6.865 | 45.81±8.065 | <0.0001 |
| Fat Mass | 18.48±6.09 | 20.96±8.18 | 20.93±10.47 | 30.46±20.58 | <0.0001 |
| HOMA1-B | 134.38±80.18 | 132.70±85.17 | 169.34±90.98 | 151.32±95.24 | 0.0429 |
| HOMA1-IR | 2.96±1.55 | 2.72±1.89 | 2.74±1.64 | 3.02±1.82 | 0.0325 |

The partial correlation analysis revealed several associations between serum uric acid (SUA) levels and various health indexes. After adjusting for age and sex, SUA showed a statistically significant positive correlation with INSULIN (FASTING) (r=0.147, p=0.0001), indicating a moderate positive association. Additionally, SUA correlated positively with BMI (r=0.044, p=0.25), Haemoglobin (HB) (r=0.066, p=0.08), and Hba1c (Glycosylated Hemoglobin) (r=0.048, p=0.21), although these associations were not statistically significant.

Conversely, SUA exhibited a significant negative correlation with Non-HDL Cholesterol (r=-0.119, p=0.002), indicating a moderate negative relationship. It also showed a significant positive correlation with Serum Creatinine (r=0.277, p<0.001), suggesting a strong positive association. Other indexes such as Mentzer Index, Platelet Count (PLT), and Serum HDL Cholesterol did not show significant correlations with SUA.

Interestingly, SUA displayed a significant negative correlation with Total Cholesterol (r=-0.138, p=0.0003), VITAMIN B12 (r=-0.130, p=0.001), and Lean Mass (r=0.099, p=0.009), indicating potential health implications related to these factors. Overall, these findings suggest a complex interplay between SUA levels and various health indexes, highlighting the need for further research to elucidate their underlying mechanisms.

### Multivariable Association Between Serum Uric Acid, Insulin Resistance, and Beta-Cell Dysfunction

In the cross-sectional analysis, the incidence of Insulin Resistance across the SUA quartiles was 69 cases (36.7%), 78 cases (42.6%), 60 cases (36.8%), and 59 cases (37.8%). The incidence of Beta-Cell Dysfunction across the SUA quartiles was 46 cases (24.5%), 38 cases (20.8%), 53 cases (32.5%), and 39 cases (25.0%). In the unadjusted model (Model 1), the ORs for Insulin Resistance were higher with increasing uric acid quartiles (p for trend < 0.001; Table 4). After adjusting for age, smoking, and alcohol consumption, the ORs and 95% CIs remained similar (Model 2). In the final model (Model 3), the magnitude of the ORs showed little change, with ORs for Insulin Resistance in the second, third, and fourth quartiles being 1.31 (1.30–1.32), 4.59 (4.57–4.61), and 2.58 (2.57–2.59), respectively (p for trend < 0.001). When SUA was included as a continuous variable, the ORs and 95% CIs for a one standard deviation increase in SUA in relation to Insulin Resistance were 1.74 (1.47–2.06). Similarly, for Beta-Cell Dysfunction, the ORs in the unadjusted model (Model 1) were slightly elevated across the quartiles (p for trend = 0.7789). After adjusting for the same covariates, the ORs for Beta-Cell Dysfunction in the second, third, and fourth quartiles were 1.59 (1.58–1.60), 0.51 (0.50–0.52), and 1.32 (1.31–1.33), respectively. The ORs and 95% CIs for a one standard deviation increase in SUA in relation to Beta-Cell Dysfunction were 0.95 (0.80–1.12), indicating no significant association.

**Table 4:** Partial Correlation Coefficients Between SUA and Other Indexes.

| Variable | Correlation Coefficient | p value |
| --- | --- | --- |
| Glucose, Fasting | 0.044049 | 0.247874 |
| HbA1c (Glycosylated Haemoglobin) | 0.047946 | 0.208434 |
| INSULIN (FASTING) | 0.147379 | 0.000102 |
| Non-HDL Cholesterol | -0.11857 | 0.00181 |
| Serum Creatinine | 0.276961 | 1.29E-13 |
| Serum HDL Cholesterol | -0.10427 | 0.006115 |
| Serum LDL Cholesterol | -0.15697 | 3.45E-05 |
| Serum Triglycerides | 0.103048 | 0.006746 |
| Serum VLDL Cholesterol | 0.086566 | 0.022963 |
| Total Cholesterol | -0.13842 | 0.000265 |
| Current Age | -0.07099 | 0.062353 |
| BMR (Basal Metabolic Rate) | 0.09066 | 0.017218 |
| BFP (Body Fat | -0.06459 | 0.090005 |
| Percentage) |  |  |
| Lean Mass | 0.099264 | 0.009077 |
| HOMA2 -B | 0.090923 | 0.016895 |
| HOMA2-IR | 0.163454 | 1.60E-05 |

**Note:** All adjusted for age and sex.
**Abbreviations:** GF, fasting glucose; HB, haemoglobin; HbA1c, glycosylated hemoglobin; INS, fasting insulin; MI, Mentzer Index; Non-HDL-C, non-HDL cholesterol; PLT, platelet count; SCr, serum creatinine; HDL-C, serum HDL cholesterol; SI, serum iron; LDL-C, serum LDL cholesterol; TG, serum triglycerides; VLDL-C, serum VLDL cholesterol; TC, total cholesterol; VB12, vitamin B12; VitD, vitamin D; Age, current age; BMR, basal metabolic rate; BFP, body fat percentage; LM, lean mass; FM, fat mass; HOMA1-B, homeostasis model assessment of beta-cell function; HOMA1-IR, homeostasis model assessment of insulin resistance.

| Condition | Model | Q1 | Q2 (OR with CI) | Q3 (OR with CI) | Q4 (OR with CI) | p for Trend |
| --- | --- | --- | --- | --- | --- | --- |
| Insulin Resistance | Model 1 | 1 | 1.11<br>(0.70–1.78) | 1.05<br>(0.65–1.70) | 1.05<br>(0.65–1.70) | 0.059 |
| Insulin Resistance | Model 2 | 1 | 0.99 (0.1–1.60) | 0.92<br>(0.56–1.50) | 0.92<br>(0.56–1.50) | 0.081 |
| Insulin Resistance | Model 3 | 1 | 1.00<br>(0.62–1.61) | 0.94<br>(0.57–1.54) | 0.94<br>(0.57–1.54) | 0.039 |
| Beta-Cell Dysfunction | Model 1 | 1 | 0.77<br>(0.46–1.29) | 0.58<br>(0.34–1.00) | 0.58<br>(0.34–1.00) | 0.231 |
| Beta-Cell Dysfunction | Model 2 | 1 | 0.75<br>(0.45–1.27) | 0.55<br>(0.31–0.97) | 0.55<br>(0.31–0.97) | 0.083 |
| Beta-Cell Dysfunction | Model 3 | 1 | 0.75<br>(0.44–1.26) | 0.54<br>(0.31–0.95) | 0.54<br>(0.31–0.95) | 0.449 |

**Notes:** 1st Quartile $\leq 282.0$ $\mu\text{mol/L}$ , 2nd Quartile = 282.1 – 334.0 $\mu\text{mol/L}$ , 3rd Quartile = 334.1 – 393.0 $\mu\text{mol/L}$ , 4th quartile $\geq 393.1$ $\mu\text{mol/L}$ . Insulin resistance was defined as HOMA-IR $\geq 2.5$ , and beta-cell dysfunction was defined as HOMA-beta within the lowest quartile among all the study participants. SUA entered the logistic regression models as quartiles or as a continuous variable. When SUA entered the model as a continuous variable, the ORs and 95% CIs for a 1 SD increase in SUA are presented. Model 1: unadjusted. Model 2: adjusted for age (continuous), drinking (yes/no), and smoking (yes/no).
**Abbreviations:** OR, odds ratio; CI, confidence interval; SUA, serum uric acid.

## Discussion

The study by [Kivity et al](https://www.liebertpub.com/doi/10.1089/jwh.2012.4043?url_ver=Z39.88-2003&rfr_id=ori%3Arid%3Acrossref.org&rfr_dat=cr_pub++0pubmed). revealed a striking gender disparity in the association between serum uric acid (SUA) and diabetes risk, with women showing a significantly stronger relationship (HR 1.57, 95% CI 1.32-1.86) compared to men (HR 1.08, 95% CI 0.99-1.17). This pronounced gender difference suggests that SUA may play a more critical role in diabetes pathogenesis in women, highlighting the need for gender-specific analyses in studies examining uric acid’s relationship to beta-cell function and insulin resistance, particularly in diverse ethnic populations.

[Cook et al](https://www.ncbi.nlm.nih.gov/pmc/articles/PMC2418956/).‘s study of 7,735 middle-aged British men revealed a complex relationship between serum uric acid and glucose levels. They found a positive correlation between uric acid and glucose up to 8.0 mmol/l, after which uric acid levels decreased. Notably, uric acid levels were significantly lower in diabetics and those with high casual glucose levels. The authors suggest that this relationship likely reflects the biochemical interaction between glucose and purine metabolism, with increased uric acid excretion during hyperglycemia and glycosuria.

[Liu et al](https://ncbi.nlm.nih.gov/pmc/articles/PMC8326926/).’s cross-sectional study of 1,230 Chinese patients with type 2 diabetes mellitus revealed a complex, nonlinear relationship between uric acid (UA) and insulin-like growth factor-1 (IGF-1). They found that IGF-1 levels increased with UA up to a threshold of 4.17 mg/dL, after which IGF-1 levels decreased as UA increased. This relationship persisted after adjusting for multiple confounding factors. The study highlights the intricate interplay between UA and IGF-1 in diabetes, suggesting that the impact of UA on IGF-1 may be concentration-dependent.

[Bhole et al](https://www.ncbi.nlm.nih.gov/pmc/articles/PMC3131180/).’s prospective study using data from two generations of the Framingham Heart Study demonstrated a strong, independent association between higher serum uric acid levels and increased risk of developing type 2 diabetes. The study, which adjusted for multiple confounding factors, found that each 1 mg/dL increase in serum uric acid was associated with a 20% higher risk of diabetes in the original cohort and a 15% higher risk in the offspring cohort. These findings suggest that elevated serum uric acid levels may be an important predictor of future diabetes risk, even in younger adults, independent of other established risk factors.

[This review](https://www.ncbi.nlm.nih.gov/pmc/articles/PMC7888074/) highlights the potential role of hyperuricemia in pancreatic β-cell dysfunction and type 2 diabetes development. Elevated uric acid (UA) levels are associated with decreased glucose-stimulated insulin secretion and β-cell death, primarily through oxidative stress and inflammation. The study proposes a threshold theory, suggesting UA’s detrimental effects occur above a specific concentration (6.7 mg/dL in rat islets). Additionally, it emphasises that UA’s effects may be potentiated by other risk factors like obesity. While the direct relationship between UA and diabetes remains controversial, emerging evidence suggests that UA-lowering drugs might be beneficial in preventing or managing diabetes, especially in at-risk individuals. However, the authors caution that more clinical trials are needed to confirm these potential benefits.

## Conclusion

## Data Availability

All data produced in the present study are available upon reasonable request to the authors

## Abstract

- Background: Brief context on hyperuricemia and its significance.
- Objective: Aim of the study.
- Methods: Overview of the study design, participants, and key methods used.
- Results: Highlight significant findings.
- Conclusions: Main conclusions and implications of the findings.

## Introduction

- **Background Information:** Define hyperuricemia and its epidemiology, particularly in the context of metabolic syndrome and diabetes.
- **Literature Review:** Summary of previous research findings linking uric acid to metabolic disturbances.
- **Study Justification:** Explanation of why this study is necessary, particularly in the Indian context.
- **Objectives:** Specific aims or hypotheses of your study.

## Methods

- **Study Design**: Describe the type of study (e.g., cross-sectional, longitudinal).
- **Participants:** Criteria for inclusion and exclusion, demographic breakdown.
- **Data Collection:** Methods used to measure serum uric acid, insulin, cholesterol levels, etc.
- **Statistical Analysis:** Description of statistical tests and models used for analysis.

## Results

- **Participant Characteristics**: Baseline characteristics of the study population.
- **Main Findings:** Results of analyses on serum uric acid levels across different quartiles, correlations with metabolic indices, etc.
- **Tables and Figures:** Include relevant visual representations of the data.

## Discussion

- **Interpretation of Results**: Discuss how your findings align or contrast with existing literature.
- **Implications:** Potential implications for clinical practice or further research.
- **Limitations**: Acknowledge any limitations of your study.
- **Future Research Directions:** Suggestions for future studies based on your findings.

## Conclusions

- Summary of Findings: Brief recap of the main findings and their significance.

## Appendices (if applicable)

- Supplementary Information Additional data, tables, or methodological details that are peripheral but relevant to the study.

## Introduction

**Hyperuricemia is a medical condition characterised by elevated levels of serum uric acid in the blood. This occurrence can stem from either an excess production of uric acid within the body or a decreased excretion of uric acid through the kidneys [1, 2]. Often underdiagnosed until later stages, hyperuricemia is prevalent in the general population, with a significant portion of affected individuals remaining asymptomatic [3]. Notably, developed countries, particularly Pacific islanders, exhibit a higher prevalence of hyperuricemia [4]**.

A recent survey conducted by the Non-Communicable Disease (NCD) department in rural India revealed a prevalence rate of 32.7% for hyperuricemia [5]. This study also noted a positive correlation between serum uric acid levels and waist circumference, along with a notable association with increased meat consumption. Further supporting this data, a meta-analysis comprising 23 studies reported a prevalence of hyperuricemia in India at 38.4% [6].

Within the Indian population, hyperuricemia is commonly associated with complications of metabolic syndrome, including Type 2 Diabetes (T2D), hypertension, and Chronic Kidney Disease (CKD) [6, 7]. Noteworthy research has highlighted hyperuricemia as an independent risk factor in the development of T2D [8, 9, 10]. Furthermore, in individuals with T2D, hyperuricemia exhibits a strong association with elevated glucose levels [11, 12].

HU has been found to be developed due to chronic insulin resistance[13]. HU seems to be associated with Insulin resistance which in turn leads to further reduced excretion of uric acid. Additionally,Uric acid levels have played a subtle role in modifying the beta cell function in men and obese patients in a Hispanic population[14]. Serum Uric Acid (SUA) levels and Homeostasis Model Assessment-Insulin Resistance (HOMA-IR) are important biomarkers for understanding the prevalence and implications of hyperuricemia in the Indian population. The prevalence of hyperuricemia varies across different regions and populations. In India, studies have reported a prevalence of around 25.8% in the general population.[15] Studies have investigated the distribution of SUA levels in the Indian population. A study published in the International Journal of Research in Medical Sciences found that the mean SUA level in a sample of 197,097 subjects was 7.45 mg/dL, with 24.66% of the subjects having hyperuricemia.[15] Another study published in the Journal of Evolutionary Medicine and Dental Sciences reported a mean SUA level of 9.32 mg/dL in rural subjects and 8.1 mg/dL in urban subjects, with a higher prevalence of hyperuricemia in rural areas.[16]

Insulin resistance is a key component of metabolic syndrome, which is associated with an increased risk of developing type 2 diabetes and cardiovascular disease. HOMA-IR is a widely used measure of insulin resistance. Research has shown that hyperuricemia is linked to insulin resistance and that elevated SUA levels are associated with increased HOMA-IR values. A study published in the Journal of Clinical and Diagnostic Research found that SUA levels were positively correlated with HOMA-IR values in a sample of Indian subjects with type 2 diabetes. [17]Another study published in the Journal of Diabetes and its Complications reported that SUA levels were higher in subjects with insulin resistance and that HOMA-IR values were positively correlated with SUA levels [18].

The available research focuses more on the relationship between SUA and insulin resistance, as measured by HOMA-IR, rather than beta-cell function (HOMA-beta) in the Indian population. There is no literature evidence on HOMA-B and SUA that we know of The prevalence of hyperuricemia is high in India, and various factors, such as diet, age, gender, and comorbidities, can influence SUA levels. However, further research is needed to specifically investigate the association between SUA and HOMA-beta in the Indian context.

**Total data -1850**

**Male data-1155**

**Hba1c (Glycosylated Hemoglobin) less than or equal to 5.7: 714**

**With homa values-328**

**Female:-258 Total 585**

